# Remote Vision Testing: Validation Of A Simple Home-Printable Vision Screening Test for Telemedicine

**DOI:** 10.1101/2020.09.01.20131698

**Authors:** Michael D Crossland, Tessa M Dekker, Joanne Hancox, Matteo Lisi, Thomas A Wemyss, Peter BM Thomas

**Author notes:** <u>Correspondence to:</u> Michael Crossland, Department of Optometry, Moorfields Eye Hospital NHS Foundation Trust, 162 City Road, London EC1V2PD.

## Abstract

**Importance:** Hundreds of thousands of ophthalmology clinic appointments have been converted to telemedicine assessments. Here we validate a simple paper vision test (the Home Acuity Test) for ophthalmology telemedicine appointments, which can be used by people who are digitally excluded.

**Objective:** To determine the repeatability of vision measured on the Home Acuity Test, and the agreement between the Home Acuity Test and the last in-clinic visual acuity.

**Design:** Bland-Altman analysis of the Home Acuity Test chart, compared to the last measured visual acuity on a standard clinical test.

**Setting:** Routine outpatient ophthalmology telemedicine clinics in a large specialist eye hospital.

**Participants:** 50 control subjects with no eye disease and 100 consecutive adult ophthalmology outpatients from strabismus and low vision telemedicine clinics. Participants were excluded if they reported subjective changes in their vision.

**Main outcomes and measures:** For control participants, test/retest variability of the HAT and agreement with standard logMAR visual acuity measurement. For ophthalmology outpatients, agreement with the last recorded clinic visual acuity and with ICD-11 visual impairment category.

**Results:** 50 control subjects and 100 ophthalmology patients with a wide range of disease were recruited. For control participants, Home Acuity Test test/retest variability was -0.012 logMAR (95% CI: -0.25 to 0.11 logMAR). Agreement with standard vision charts was -0.14 logMAR, with a 95% confidence interval of -0.39 to +0.12 logMAR. For ophthalmology outpatients, agreement in visual acuity was -0.10 logMAR (one line on a conventional logMAR sight chart), with the Home Acuity Test indicating poorer vision than the previous in-clinic test. The 95% confidence interval for difference was –0.44 to +0.24 logMAR. Agreement in visual impairment category was good for patients (Cohen’s *κ* test, *κ =* 0.77 (95% CI, 0.74 to 0.81), and control participants (Cohen’s *κ* test, *κ =* 0.88 (95% CI, 0.88 to 0.88).

**Conclusions and relevance:** The Home Acuity Test can be used to measure vision by telephone for a wide range of ophthalmology outpatients with diverse conditions, including those who are severely visually impaired. Test/retest variability is low and agreement in visual impairment category is good.

---

Eye problems are the cause of about 1.5% of all consultations with family doctors^1^ and ophthalmology is the largest specialty in the English healthcare system, with nearly 8 million ophthalmology outpatient appointments taking place in England every year.^2^ In response to the COVID-19 outbreak, in March 2020 the UK’s Royal College of Ophthalmologists recommended that ‘all face-to-face outpatient activity should be postponed unless patients are at high risk of rapid, significant harm if their appointment is delayed’.^3^ Over the next three months more than 100,000 ophthalmology outpatient appointments were cancelled at Moorfields Eye Hospital alone.

Video and telephone-based consultations have been used successfully in ophthalmology,e,g.^4-6^ but measuring vision remotely remains a challenge. Computer-based assessments of vision require careful calibration of viewing distance, screen size and screen luminance.^7^ Trials of computer based vision measurements have involved giving participants laptops with known screen parameters.^8,9^

Smartphones can be used to measure visual acuity,^10^ but a 2015 review of smartphone vision testing apps found poor agreement with conventional vision tests, particularly for people with severe vision impairment.^11^ Further, more than 1 in 8 people in the UK do not use the internet, including 48% of those over 75 years of age,^12^ all of whom are classified as being of at least moderate risk of complications from Covid-19.^13^ People with lower household income are also more likely to be digitally excluded.^12^

These difficulties have caused ophthalmology services to ask patients to judge their performance on daily living tasks such as reading television subtitles, to measure certain aspects of vision but not distance visual acuity, to defer visual acuity testing completely, or to use printed charts.^14,15^

Organisations such as the American Academy of Ophthalmology^16^ and the UK’s College of Optometrists^17^ have made paper vision charts to be downloaded and printed. These tests do not meet the international guidelines for progression of letter size,^18,19^ do not have crowding bars, and have not been validated. Further, only one chart is available from each organisation, making repeat measurement inaccurate: people are likely to remember the letters when reading with their second eye.

Here we assess the Home Acuity Test (HAT, available at homeacuitytest.org), a simple, open source, vision screening test which can be printed on A4 paper and posted to patients. It could also be downloaded and printed by patients for urgent or emergency assessment – a silhouette of a credit card is included with the downloadable test to ensure that the printed size of the image is correct, and that no expected scaling has taken place. It incorporates a logarithmic scaling of letter size and crowding bars. Several billion^*^ unique charts can be downloaded from homeacuitytest.org for home monitoring of vision.

## Method

The home acuity test (HAT) was developed by the authors to measure visual acuity remotely for those unable to attend hospital ophthalmology appointments due to the COVID-19 outbreak.

The HAT chart consists of 18 randomly selected Sloan letters^20^ displayed over five lines. Letter size progresses logarithmically, with letters being half the size of those on the preceding line. Four letters are presented on each line except the first line, which contains two letters. Crowding bars of one stroke width surround each line of letters, with a one-stroke-width gap between the letter edge and the crowding bar.

From 150cm, the largest letters subtend 1.3 logMAR (3/60) and the smallest line measures 0.1 logMAR (6/7.5).

### Control subjects

Fifty adults were recruited from Moorfields Eye Hospital staff. All had good corrected vision (of at least 0.1 logMAR (6/7.5) with spectacles). Participants covered their better eye with their hand and read two versions of the HAT chart from 150cm with their poorer eye. Spectacles were not worn, to avoid a ceiling effect and to determine whether the HAT overestimated vision for those with uncorrected myopia, due to the chart’s closer viewing distance. Data were recorded in two ways: as a total number of letters read, and using a line assignment method where credit was given for a line where half of the letters were read correctly.

Participants also read a conventional retroilluminated ETDRS logMAR visual acuity chart from the standard distance of 4 metres, with data recorded using letter-by-letter scoring.

Bland-Altman analysis determined agreement between vision measured using the HAT and the conventional logMAR chart, and test-retest variability of the HAT.^21^

### Ophthalmology outpatients

Since early April 2020, HAT charts have been posted to outpatients having telephone consultations in two adult ophthalmology clinics as part of a service improvement project: strabismus, and low vision. Patients were sent two HAT charts along with Blue-Tac to fix charts to a wall and a 150cm length of string to measure chart viewing distance. Patients were phoned by a clinician (optometrist, orthoptist or ophthalmologist). As part of their telephone assessment, participants were asked to cover each eye in turn and to read the HAT from 150cm, using distance glasses if necessary. The number of letters read correctly was recorded.

Data were analysed for one hundred patients who self-described their vision as stable. Data were analysed from the right eye only, unless visual acuity was not recordable from the right eye (i.e. visual acuity was poorer than 1.3 logMAR (3/60)) in which case the left eye was used. Bland-Altman analyses were used to compare home-measured vision on the HAT to the last visual acuity value measured in an ophthalmology clinic.

For ophthalmology patients, vision was classified into categories of visual impairment loosely based on the International Classification of Disease-11 guidelines (ICD-11,^22^ table 1). Exact agreement with the WHO guidelines was not possible due to the levels of vision measured on the HAT.

**Table 1.** Categorisation of visual impairment by HAT, and ICD-11 guidance.

|  | **HAT categorisation** | **ICD-11 categorisation** |
| --- | --- | --- |
| No visual impairment | 0.1 logMAR | 0.3 logMAR or better |
| Mild visual impairment | 0.4 logMAR | Worse than 0.3 logMAR |
| Moderate visual impairment | 0.7 logMAR | Worse than 0.5 logMAR |
| Severe visual impairment | 1.0 logMAR | Worse than 1.0 logMAR |
| Blind | 1.3 logMAR | Worse than 1.3 logMAR |

### Ethical approval and funding

The study of control subjects was approved by the UCL Research Ethics Committee, code 4846/001. Informed consent was collected from all participants.

Approval for the retrospective analysis of patient data was obtained from the Clinical Audit committee at Moorfields Eye Hospital NHS Foundation Trust (service improvement approval number 619).

The authors affirm that the manuscript is an honest, accurate, and transparent account of the study being reported; that no important aspects of the study have been omitted; and that any discrepancies from the study as originally planned have been explained.

No external funding was received for this study. The HAT is open source (https://github.com/twemyss/hat) and free of charge, and the authors have no commercial interest in the test.

## Results

### Control subjects

The mean age of participants was 36 years (sd: 10.8 years, range 22-61 years). Thirty-three (66%) were female. Mean vision in the tested eye was 0.15 logMAR (6/7.5-; sd: 0.39, range -0.3 to 1.1 logMAR).

#### Agreement between HAT and conventional logMAR chart

Mean difference in VA was -0.16 logMAR (less than 2 lines on a conventional logMAR sight chart), with the clinic chart indicating better vision than the HAT. The 95% confidence interval for the difference between the HAT and the conventional chart was –0.42 to +0.09 logMAR (figure 1).

**Figure 1.**
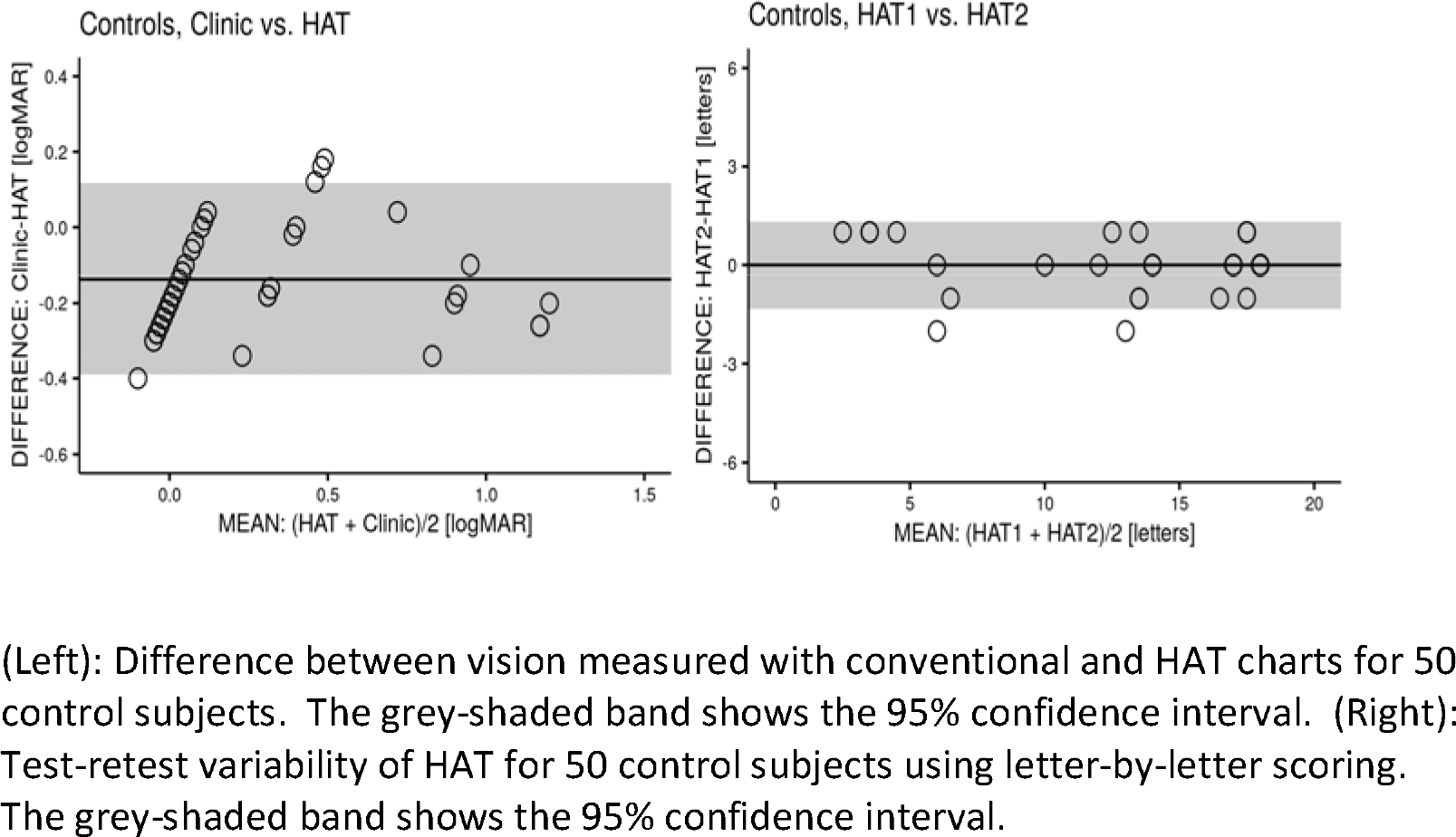
(Left): Difference between vision measured with conventional and HAT charts for 50 control subjects. The grey-shaded band shows the 95% confidence interval. (Right): Test-retest variability of HAT for 50 control subjects using letter-by-letter scoring. The grey-shaded band shows the 95% confidence interval.

The best visual acuity measurable on the HAT is 0.1 logMAR (6/7.5). Post-hoc evaluation showed that 31 participants had better vision than this on the conventional chart. In effect these values are censored: the true acuity may be better than 0.1 but it is unknown whether this is the case, or by how much. To account for this, a further Bland-Altman analysis was performed using maximum likelihood estimation for censored data.^23^ This revealed a mean difference between charts of -0.14 logMAR, with a 95% confidence interval of -0.39 to +0.12 logMAR (figure 1).

#### Effect of uncorrected myopia

Thirteen participants (26%, with a mean age of 34 years) had myopia and read the chart without spectacles. For this subgroup, the mean difference in vision measured with HAT and conventional charts was -0.09 logMAR (sd: 0.15; range -0.34 to +0.18 logMAR), indicating that visual acuity was about 1 line worse on the HAT than on the conventional chart. The closer test distance of the HAT did not overestimate visual acuity.

#### Repeatability of HAT

Using letter-by-letter scoring, the mean difference in VA between two versions of the HAT was 0 letters (sd: 0.67 letters, range -2 to +1 letters; figure 2). When using line assignment scoring the mean difference was -0.012 logMAR (sd: 0.06; range -0.3 to 0 logMAR).

**Figure 2.**
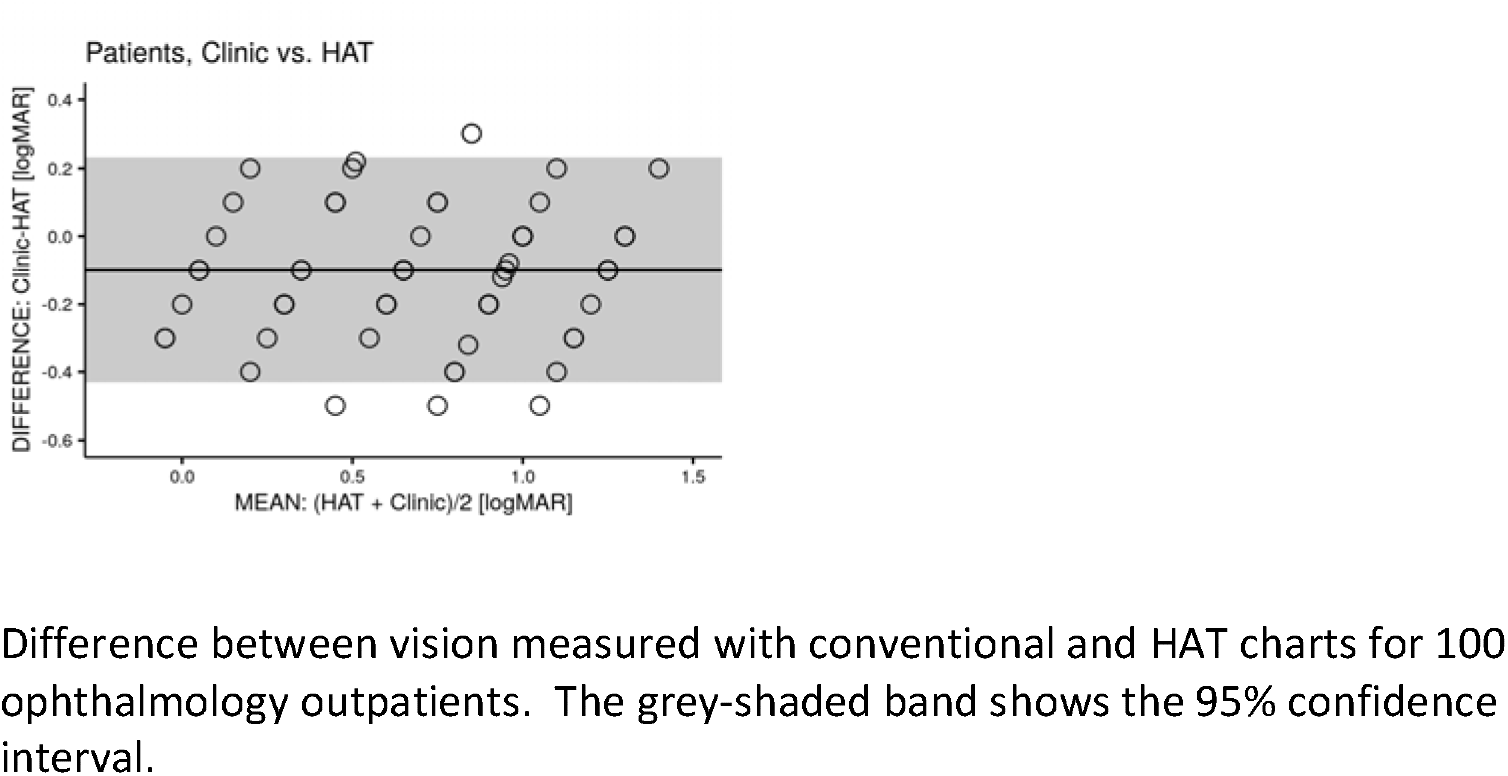
Difference between vision measured with conventional and HAT charts for 100 ophthalmology outpatients. The grey-shaded band shows the 95% confidence interval.

### Ophthalmology outpatients

Mean age was 55 years (sd: 22.2 years, range: 16 – 97 years). Sixty-five were female. Primary diagnosis was retinal disease in 24 participants, age-related macular degeneration in 14 and strabismus in 14, macular dystrophy in 10, glaucoma in 8, neuro-ophthalmological disease in 8, and others in 22 participants. Mean visual acuity in the test eye was 0.78 logMAR (sd: 0.37, range: 0.1 – 1.3 logMAR) by HAT.

#### Agreement between HAT and last clinic VA

Mean difference in VA was -0.10 logMAR (one line on a conventional logMAR sight chart), with the HAT indicating poorer vision than the previous in-clinic test. The 95% confidence interval for difference was –0.44 to +0.24 logMAR (figure 3).

**Figure 3.**
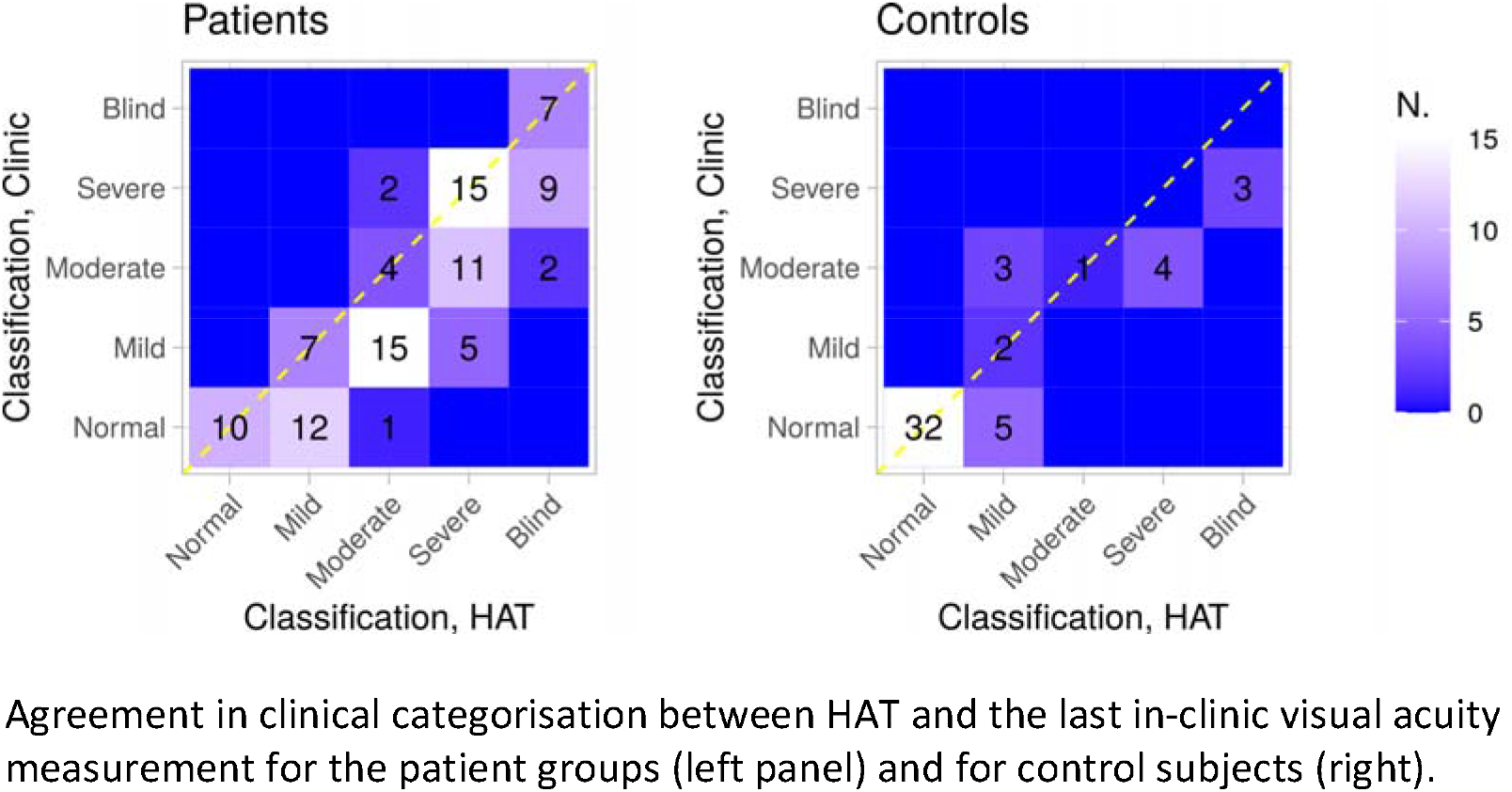
Agreement in clinical categorisation between HAT and the last in-clinic visual acuity measurement for the patient groups (left panel) and for control subjects (right).

HAT classified 10 patients as not visually impaired, 19 as having mild visual impairment, 22 as moderately visually impaired, 31 as severely visually impaired and 18 as blind. The agreement between the categorical classification resulting from the in-clinic and the HAT tests was quantified using Cohen’s *κ*, which revealed good agreement between the two, *κ =* .77 (95% CI, .74 to .81, Figure 4).^24^ The same analysis for control participants showed an agreement of *κ =* 0.88 (95% CI, 0.88 to 0.88). Figures 4 and 5 show agreement in visual impairment classification between HAT and the last clinic test, for both patients and control group.

**Figure 4.**
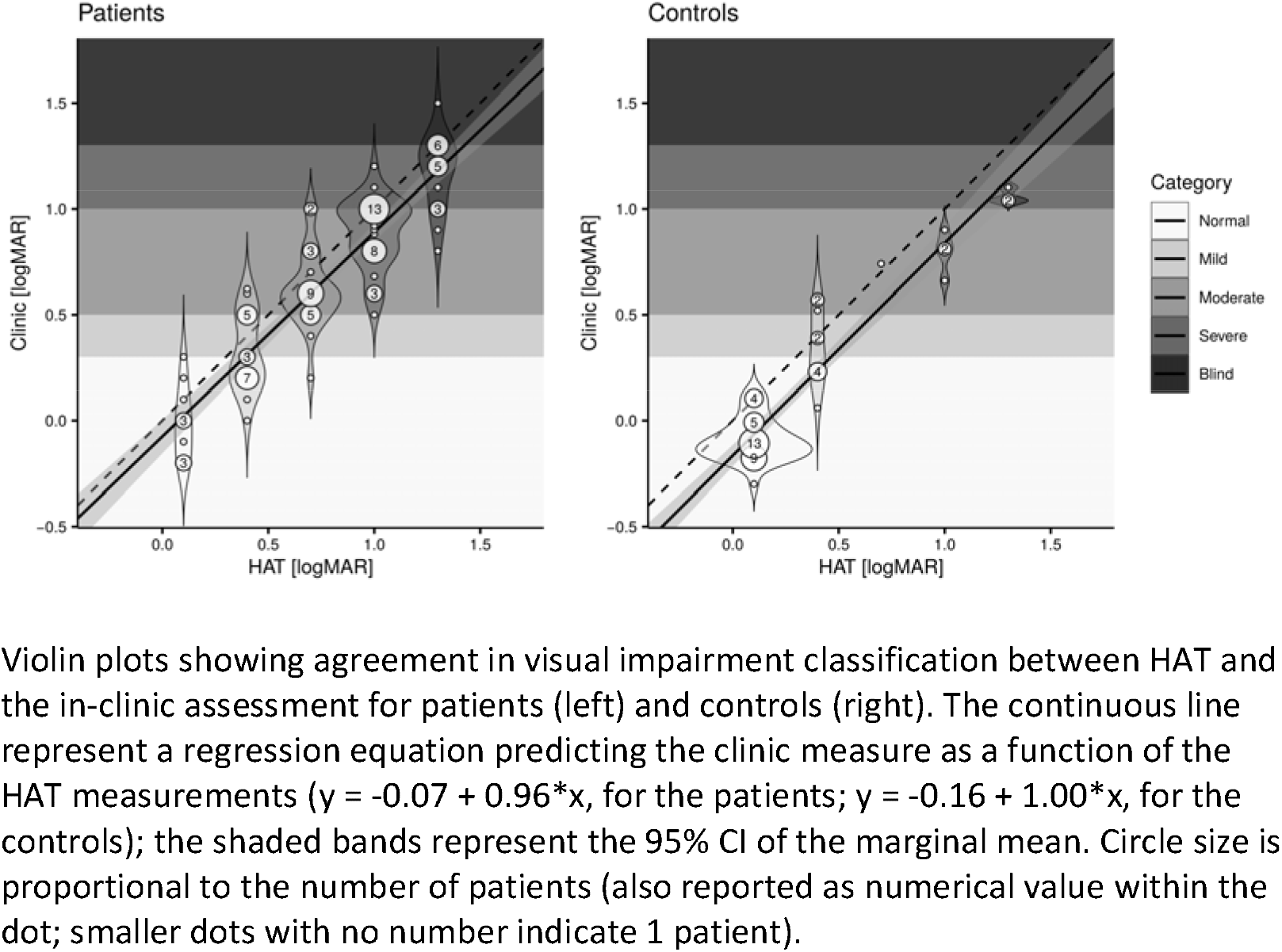
Violin plots showing agreement in visual impairment classification between HAT and the in-clinic assessment for patients (left) and controls (right). The continuous line represent a regression equation predicting the clinic measure as a function of the HAT measurements (y = -0.07 + 0.96*x, for the patients; y = -0.16 + 1.00*x, for the controls); the shaded bands represent the 95% CI of the marginal mean. Circle size is proportional to the number of patients (also reported as numerical value within the dot; smaller dots with no number indicate 1 patient).

## Discussion

We have shown that vision can be measured using a simple printed vision test. Test repeatability is high and compares very favourably to data published for conventional and computer based clinic charts.^25 26^ Mean agreement with standard clinical tests was within 5 letters for patients and control participants.

The high test-retest variability of the HAT should give confidence in identifying new intraocular differences in vision, making it particularly appropriate for monitoring vision in people with strabismus, or with disease likely to worsen in one eye. Although we have only presented data for a letter chart, the HAT website includes a symbol version of the test is available for children and those who can not identify letters, using the Auckland optotypes.^27^

Asking patients to self-report their vision on a printed test in their own home differs from measuring visual acuity in a clinic in several important ways. First, the contrast of the printed chart is less than a conventional logMAR chart. Contrast sensitivity is reduced in people with a wide range of eye diseases.^28,29^ Second, lighting at home is likely to be far lower than a typical hospital clinic, by as much as ten times.^30^ Third, patients may have been confused about the exact instructions for reading a chart at home: for example, they may have covered the wrong eye, been wearing the wrong spectacles, or been standing at an incorrect distance. Fourth, their motivation may have been lower for a home based assessment.

Despite these important differences we found good agreement between the HAT and the last in-clinic visual acuity measurement. The mean difference in acuity was just over one line of visual acuity. Where differences existed, the HAT erred on the side of safety, tending to underestimate vision compared to the last clinical test. As figure 4 shows, only 2% of patients were categorised as having better vision by home testing.

Although our patients self-reported stable vision, their sight may have deteriorated since their previous assessment. The mean time between the last clinic appointment and the HAT assessment was 11.6 months (range 1-69 months). There was no relationship between the time since the previous in-clinic vision measurement and the difference between HAT and in-clinic vision measurements (linear regression, r=0.067). When routine ophthalmology clinics are reopened, direct comparison of the HAT and a standard clinical chart can be performed on the same day. This will help to determine if the difference in performance between HAT and LogMAR tests of vision are due to features of the test, such as its granularity (acuity lines are separated by 0.3 logMAR) or difference in testing circumstances (e.g. encouragement from a clinic nurse, lighting conditions).

Unlike the AAO and COptom tests, our design has a geometric progression of letter sizes, as advocated by the European Standard and the International Council of Ophthalmology.^18,19^ This design principle allows testing at any distance: for example, extending the test distance to 190cm allows visual acuity to be measured between 1.0 logMAR (6/60) and 0.0 logMAR (6/6). This would reduce the impact of having an upper acuity limit of 0.1 logMAR for those with good vision, for example in a routine optometry clinic or a refractive surgery service.

Our charts also include crowding bars. Without crowding bars, letters in the middle of a line are more difficult to identify than the first or last letter on a row, and noncrowded letters overestimate visual acuity in people with many eye conditions, including macular degeneration,^30^ amblyopia,^32^ and nystagmus.^33^

Measuring vision at home is unlikely to ever be as accurate as in-clinic assessment by a trained clinician, but we show that the HAT could become an objective measure of visual performance in people with and without eye disease. By avoiding digital devices, it is likely to be accessible to more of the population, and less liable to unexpected variation in performance.

## Data Availability

complete de-identified data is available from the corresponding author

https://github.com/twemyss/hat

## Acknowledgements

We thank Katy Barnard, lead optometrist in the low vision clinic, and all clinical staff who performed vision measurements by telephone or provided feedback on earlier iterations of this test.

## Footnotes

* There are actually 58 quadrillion possible values for the chart: to be exact 58,071,715,430,400,000

